# A composite endpoint for acceptability evaluation of oral drug formulations in the pediatric population

**DOI:** 10.1101/2021.08.20.21262333

**Authors:** Manfred Wargenau, Sibylle Reidemeister, Ingrid Klingmann, Viviane Klingmann

**Author notes:** **Corresponding author:** Viviane Klingmann, MD, PhD.

## Abstract

**Introduction:** A medicine’s acceptability is likely to have significant impact on pediatric compliance. EMA and FDA guidance on this topic ask for investigation of acceptability. Although palatability and deglutition are denoted as elements of acceptability, the impact of both on acceptability remains unclear as an unambiguous definition of acceptability is lacking. Actually, globally applied standards for acceptability definition, testing methodology and assessment criteria do not exist. A definition of acceptability establishing a composite endpoint that combines deglutition and palatability in different age groups is presented here.

**Methods:** This composite acceptability endpoint is based on validated assessment methods for deglutition and palatability in children of different age groups with different galenic placebo formulations, in line with criteria EMA proposed for assessing acceptability in children from newborn to 18 years. Data from two studies investigating mini-tablets, oblong tablets, orodispersible films and syrup were used to investigate the validity, expediency and applicability of the suggested composite acceptability assessment tool.

**Results:** The new composite endpoint is highly suitable and efficient to distinguish preferences of oral formulations: Mini-tablets and oblong tablets were significantly better accepted than syrup and orodispersible film.

**Conclusion:** Since the suggested acceptability criterion takes both deglutition and palatability into account as composite endpoint, it is highly sensitive to detect acceptability differences between oral formulations. It is a well-defined, valid approach, which particularly meets regulatory requirements in an appropriate and comprehensive manner and may in future serve as an easy, standardized method to assess and compare acceptability of pediatric formulations with active substances.

## Introduction

The oral route of administration is the most commonly used for pediatric medicines, which are developed according to ethical obligations and regulatory guidelines. The FDA Draft guidance [1] generally asks for “the ethical acceptability”, and the EMA guideline [2] states that “… at least considerations for the choice of route(s) of administration, dosage form(s), dosing needs/flexibility and excipients in the preparation and administration device(s) should be discussed, taking into consideration acceptability”. In particular, the following aspects are to be considered when selecting an age-appropriate formulation: “The patient acceptability including palatability (e.g., local pain, taste)”. Furthermore, this guideline mentions in section 10 another characteristic of acceptability, namely swallowability.

Given the increasing focus to develop patient centric formulations as of today, existing acceptability assessment methodology and approaches are fragmented [3,4], and a harmonized approach is lacking [4]. Actually, comprehensively investigated and globally established standards for acceptability definition, test methods and assessment criteria do not exist: in most published studies the results were based on surveys or observations by parents, cares, healthcare providers or patients, or on underpowered studies with different assessment conditions. In 2013, Klingmann et al. published the results of a standardized, in-person controlled study with mini-tablets and syrup with a statistically defined sample size and defined scores for acceptability and swallowability observed and documented by a trained investigator [6]. In further statistically powered studies this validated method was applied to investigate acceptability of multiple mini-tablets, orodispersible films and oblong tablets [5-10]. However, although palatability and swallowability assessing the act of deglutition are denoted as elements of acceptability, the relationship between these key terms remains unclear.

This project is intended to establish and validate an acceptability test system as a composite endpoint based on deglutition and palatability, for which established, broadly accepted definitions are used (Klingmann et al. [5-10]) in boys and girls from newborn to 18 years across different races comparing four oral pediatric drug formulations: syrup, mini-tablets, capsules and tablets. It is intended to broadly discuss the results with academic and industry experts, clinicians, regulators and patients to provide an internationally accepted method for acceptability assessment. In addition, the suggested acceptability assessment procedure may in future serve as a test system to enable patient centric drug development [11].

## Materials and Methods

According to the validated method of Klingmann [12] in children between 2 days and 6 years acceptability can be assessed by observing the act of deglutition and a rapid mouth inspection by a trained investigator. The outcome of the deglutition was described according to the following scoring scheme:

A drug formulation was considered as “acceptable” when it was either “completely swallowed” or “partially swallowed”.

Palatability can be described as an expression in mimic, gestures and – in older children - expressed opinion - of an overall appreciation of an oral medicine towards its appearance, smell, taste, after taste and mouth feel (e.g. texture, cooling, heating, trigeminal response) [13].

In two studies in children from newborn to 6 years, Klingmann et al. [9] and Münch et al. [10] validated a method that was based on assessing the palatability by video documentation and independent evaluation by two blinded raters according to the following scoring scheme:

The palatability assessments of the two raters are combined according to the following rule:

Assuming that a combination of deglutition and palatability would describe acceptability more precisely, the composite endpoint was developed defining acceptability as ‘high’, ‘good’, ‘low’, or ‘no’ based on deglutition and combined rater palatability as shown in the following table:

The validity of this combined criterion for acceptability has been investigated by applying factor analysis. The following variables have been submitted to factor analysis:

- deglutition score (1, 2, 3, 4, 5), refer to table 1
- palatability score (1, 2, 3) for rater 1, refer to table 2
- palatability score (1, 2, 3) for rater 2, refer to table 2.

**Table 1:** Scoring criteria for deglutition

| Score | Observation |
| --- | --- |
| 1 | Completely swallowed |
| 2 | Partially swallowed<br>(chewed and/or parts of the solids or syrup were found during oral inspection) |
| 3 | Spat out |
| 4 | Swallowed the wrong way<br>(cough may have been caused) |
| 5 | Refused to take |

**Table 2:** Palatability scoring criteria based on video documentation per rater

| Score | Assessment | Interpretation |
| --- | --- | --- |
| 1 | Pleasant | Positive hedonic pattern:<br>Tongue protrusion, smack of mouth and lips, finger sucking, corner elevation |
| 2 | Neutral | Neutral mouth & body movements, and face expression |
| 3 | Unpleasant | Negative aversive pattern:<br>Gape, nose wrinkle, eye squinch, frown, grimace, head shake, arm flail |

**Table 3:**
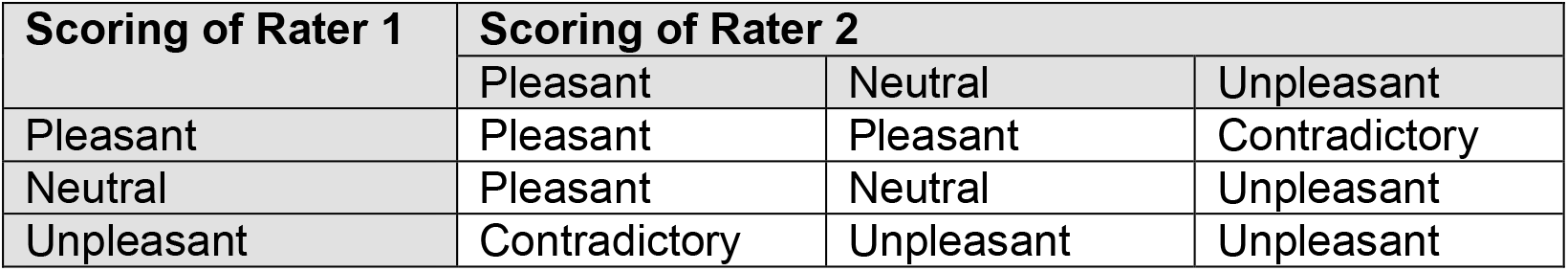
Combined rater palatability assessment

## Results

Evaluation and validation of acceptability as composite endpoint was performed using the data from two previous studies:

Study_1: “Acceptability of small-sized oblong tablets in comparison to syrup and mini-tablets in infants and toddlers: A randomized controlled trial”, Münch et al. [10]. In total, 280 children stratified into 5 age groups were included (1 to <2 years, 2 to <3 years, 3 to <4 years, 4 to <5 years, 5 to <6 years).

Study_2: “Acceptability of an orodispersible film compared to syrup in neonates and infants: A randomized controlled trial”, Klingmann et al. [9]. In total, 150 children stratified into 3 age groups were included (2 to 28 days, 29 days to 5 months, > 5 to 12 months).

### Factor analysis

Factor analysis was separately applied for each formulation, i. e. for syrup, oblong tablets and mini-tablets from Study_1, and for syrup and orodispersible film from Study_2.

Deglutition, palatability (reader1) and palatability (reader2) were included in the factor analyses. Here, results are exemplarily given for the syrup formulation (Study_1) since it has been widely used and therefore represents an appropriate reference formulation:

The correlation between the three assessments ranged between 0.684 and 0.777 and resulted in a high value of 0.891 for Cronbach’s standardized alpha. Factor analysis clearly identified one main component with an eigenvalue of 2.32 (presenting a portion of 77%), other eigenvalues were clearly below 1 with 0.46 and 0.22. Thus, it can be concluded that one dominant main factor exists comprising the information from the three single assessments, and this condensed information can be interpreted as ‘acceptability’.

Factor loads for deglutition, palatability (reader1) and palatability (reader2) were found comparable with values of 0.82, 0.91, and 0.90, thus contributing to a similar extent to the main component. This can be presented as linear combination of the single variables by using the above-mentioned factor loads.

The results for acceptability defined as composite endpoint according to table 4 were calculated for the syrup formulation. Each outcome category of acceptability was then related to the outcome of the factor analysis as expressed by the linear combination for the main component (Table 5).

**Table 4:** Assessment of acceptability as composite endpoint

| Palatability | Deglutition Score |  |  |
| --- | --- | --- | --- |
| | 1 | 2 | $\geq 3$ |
| Pleasant | High | Good | No |
| Neutral | Good | Low | No |
| Unpleasant | Low | No | No |
| Contradictory | Good | Low | No |

**Table 5:** Relationship between acceptability rates and main component derived from factor analysis (N = 141) for syrup formulation from Study_1

| Acceptability (composite endpoint) | n (%) | Mean (SD) of main component (linear combination) |
| --- | --- | --- |
| High | 39 (27.7%) | 3.21 (0.438) |
| Good | 27 (19.2%) | 4.44 (0.017) |
| Low | 12 (8.5%) | 5.79 (0.483) |
| No | 63 (44.7%) | 7.20 (0.636) |

**Table 6:** Deglutition results for different formulations, ODF: orodispersible film

| Deglutition | Study_1 |  |  | Study_2 |  |
| --- | --- | --- | --- | --- | --- |
|  | Syrup | Mini-tablet | Oblong tablet | Syrup | ODF |
|  | n (%) | n (%) | n (%) | n (%) | n (%) |
| Completely swallowed | 76 (53.5%) | 113 (80.4%) | 215 (76.0%) | 73 (48.7%) | 105 (70.0%) |
| Partially swallowed | 38 (26.8%) | 10 (7.1%) | 24 (8.5%) | 48 (32.0%) | 38 (25.3%) |
| Spat out | 5 (3.5%) | 5 (3.6%) | 15 (5.5%) | 20 (13.3%) | 0 (0%) |
| Swallowed the wrong way | 0 (0%) | 0 (0%) | 0 (0%) | 2 (1.3%) | 0 (0%) |
| Refused to take | 23 (16.2%) | 13 (9.2%) | 29 (10.3%) | 7 (4.7%) | 7 (4.7%) |
| Total | 142 (100%) | 141 (100%) | 283 (100%) | 150 (100%) | 150 (100%) |

**Table 7:** Acceptability results as composite endpoint for different formulations, ODF: orodispersible film

| Acceptability | Study_1 |  |  | Study_2 |  |
| --- | --- | --- | --- | --- | --- |
|  | Syrup | Mini-tablet | Oblong tablet | Syrup | ODF |
|  | n (%) | n (%) | n (%) | n (%) | n (%) |
| High | 39 (27.7%) | 68 (48.6%) | 121 (43.5%) | 28 (18.7%) | 22 (15.1%) |
| Good | 27 (19.2%) | 47 (33.6%) | 98 (35.3%) | 49 (32.7%) | 59 (40.4%) |
| Low | 12 (8.5%) | 4 (2.9%) | 12 (4.3%) | 27 (18.0%) | 47 (32.2%) |
| No | 63 (44.7%) | 21 (15.0%) | 47 (16.9%) | 46 (30.7%) | 18 (12.3%) |
| Total | 141 (100%) | 140 (100%) | 278 (100%) | 150 (100%) | 146 (100%) |

A high association between acceptability categories and the results from factor analysis can be recognized. Comparison of the acceptability categories with regard to the main component by analysis of variance yielded a p-value < 0.0001.

Thus, the suggested acceptability as composite endpoint can be regarded as a valid approach representing the result of the factor analysis.

Factor analyses were analogously performed for the other 4 formulations (oblong tablets and mini-tablets from Study_1, and for syrup and orodispersible film from Study_2). In all cases highly consistent results to those presented above for syrup (Study_1) were obtained, thus providing high validity and reliability of the suggested approach for assessing acceptability as composite endpoint.

### Application of the acceptability approach as composite endpoint

The two studies considered in this work used deglutition as single criterion for acceptability, the respective results are presented below:

Results concerning good and high acceptability are graphically displayed in Figure 1.

**Figure 1:**
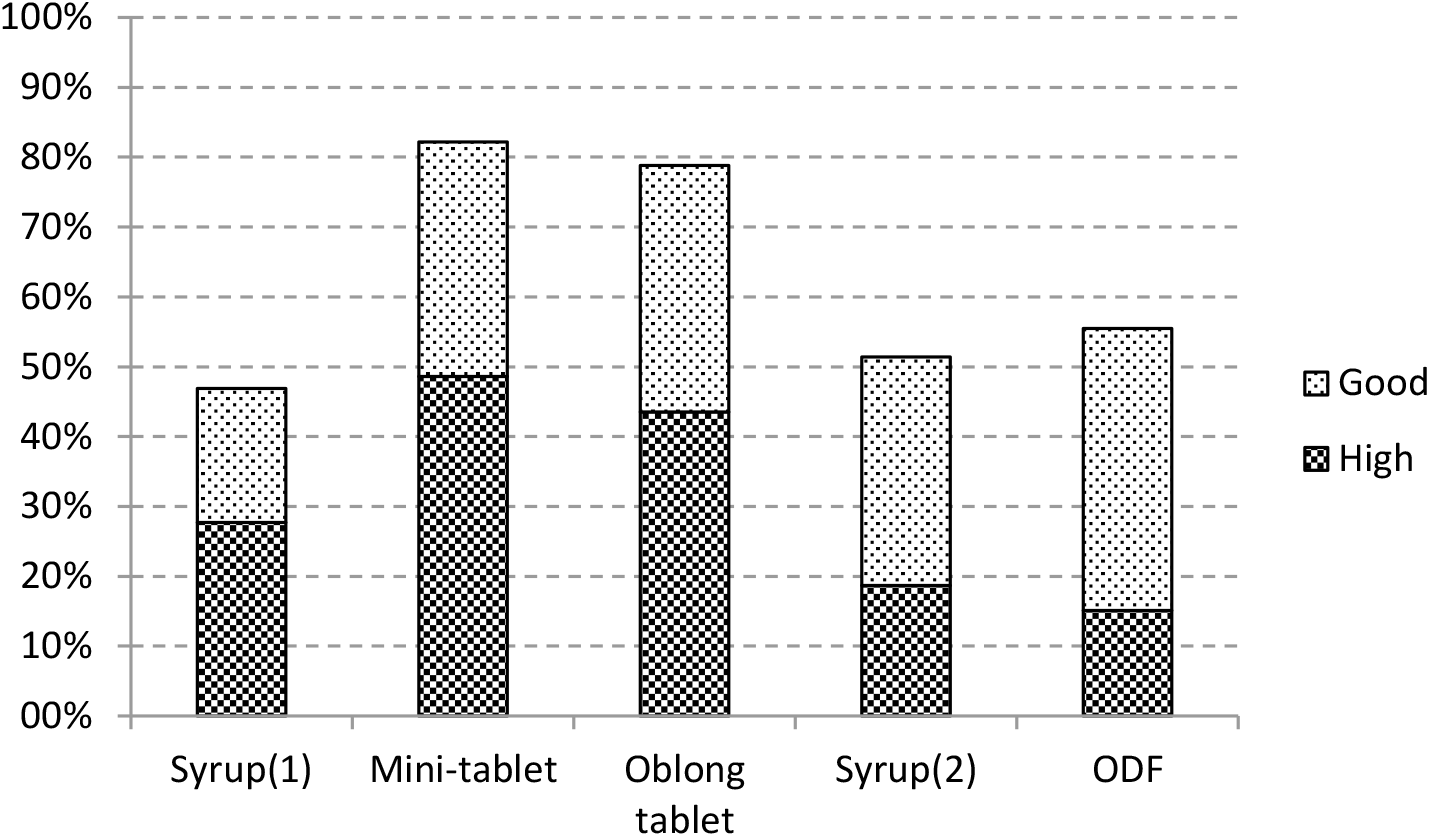
Results of good or high acceptability as composite endpoint for different formulations, ODF: orodispersible film

Mini-tablet and oblong tablet show much better results compared to syrup and ODF when considering acceptability as ‘good or high’. ‘High’ acceptability is clearly observed at higher rates for mini-tablet and oblong tablet compared to the other formulations.

The outcome of the newly developed acceptability definition as composite endpoint (regarding ‘good or high’ acceptability) is compared to the previously used definition of acceptability which was based on swallowability assessments only (refer to Figure 2).

**Figure 2:**
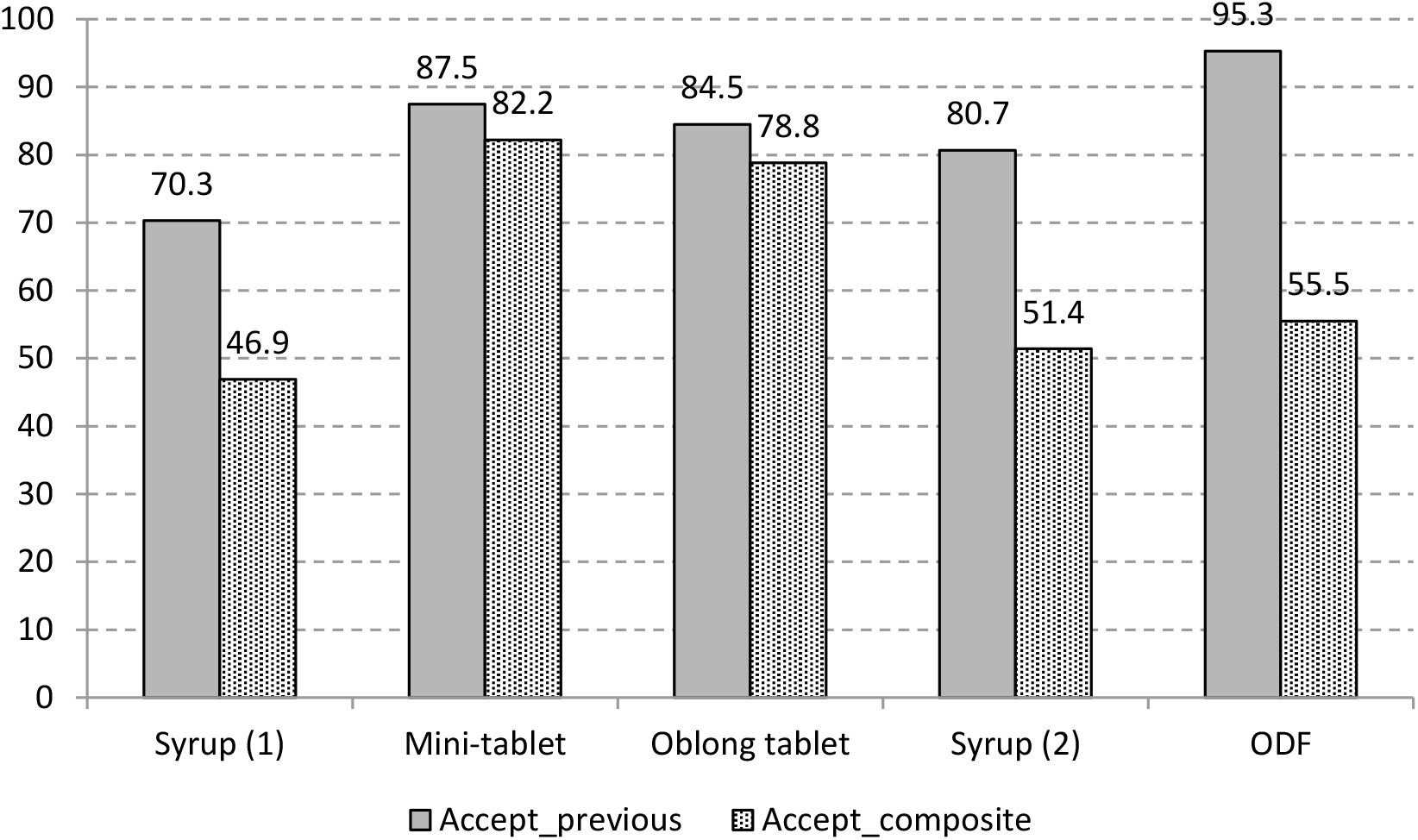
Comparison of results of different definitions of acceptability: Accept_previous: acceptability solely based on deglutition, Accept_composite: acceptability as composite endpoint, ODF: orodispersible film

The newly defined composite endpoint method discriminates more between the four different formulation principles than the previous definition which was based on deglutition only: solid dosage forms, e.g., mini tablets and oblong tablets show higher acceptability over a liquid dosage form, e.g., syrup in children aged 2 days to 6 years.

## Discussion

Starting in 2009 with a first prospective uncontrolled, single-dose study with a single 3 mm mini-tablets in 100 children aged 2 to 6 years, Thomson et al. [14] used an observation score distinguishing between “swallowed”,” chewed”, “spat out” or “refused to take”. In further studies by other authors assessment methods included observations by parents, cares, patients with different assessment scores based on opinions or observation, visual analogue scales, or by trained investigators under highly standardized conditions and video observation [4]. Different parameters were assessed like acceptability, swallowability and palatability after formulation administration occurred with different vehicles like drinks or soft food.

Similar diverse attempts were made to investigate multiple mini-tablets and other oral galenic formulations like orodispersible films, tablets, oblong tablets, capsules, sprinkles, etc. The results of these studies are of different reliability and comparability [4].

Due to the lack of a validated approach, assessing and comparing acceptability of pediatric solid oral dosage forms is currently not possible [11]. This gap is intended to be closed with the proposed composite endpoint based on deglutition and palatability. Its suitability was demonstrated by evaluating data of two published studies [9,10] in children aged newborn to 6 years. The data of the underlying studies were based on validated, standardized assessment methods and followed the existing regulatory guidance’s and requirements. The composite endpoint clearly increased the differentiation of acceptability for different pediatric oral formulations. Since previous studies revealed sufficiently large inter-rater reliability, palatability as one component of the combined endpoint could also be assessed by only one rater of a video or other assessment methods like observation by a second investigator or facial hedonic scale in older children. To ensure the suitability of this composite endpoint for all age groups and for different oral formulations, a planned confirmatory, statistically powered study shall provide the evidence.

In summary, since the acceptability assessed as composite endpoint from standardized measurement procedures takes both - deglutition and palatability - into account, it is highly sensitive to detect differences between formulations. It is a well-defined and valid approach which appropriately and comprehensively meets regulatory requirements and is easy to apply in active pharmaceutical ingredients trial. The suitability of this composite endpoint should be broadly discussed to potentially enable alignment of competent authorities, sponsors and clinicians about the judgement on acceptability of pediatric solid oral formulations.

Especially with the application of the composite endpoint for acceptability in the reevaluation of the two studies it becomes obvious that the already started paradigm shift from oral liquid formulations to solid oral formulations in young children will lead to more patient-centric approaches in the development of better medicines for children.

## Conclusions

With this composite endpoint a suitable, easy to apply, guideline-conform method to assess acceptability based on deglutition and palatability was established. This method is able to reliably detect differences between pediatric oral formulations, and may in future serve as a test system to enable patient centric drug development in pediatric populations.

## Supporting information

CONSORT Checklist

Disclosure form

## Data Availability

Data used to calculate the composite endoint in this article is already published in several papers.

## Funding statement

None

## Conflict of interest statement

No conflict of interest

## Author contributions

Manfred Wargenau contributed substantially to the conceptualization, design and methodology of the article, to the data curation as well as formal analysis. He participated in the writing of the initial draft as well as in review and editing of the manuscript. Sibylle Reidemeister contributed substantially to the conceptualization, design and methodology of the article, as well as the funding acquisition. She participated in the writing of the initial draft as well as in review and editing of the manuscript. Ingrid Klingmann contributed substantially to the conceptualization, design and methodology of the article. She participated in the writing of the initial draft as well as in review and editing of the manuscript. Viviane Klingmann contributed substantially to the conceptualization, design and methodology of the article and had the oversight of the project. She participated in the writing of the initial draft as well as in review and editing of the manuscript.

## Notes

### Competing Interest Statement

The authors have declared no competing interest.

### Funding Statement

No funding.

### Author Declarations

No approval was necessary.

xI have followed all appropriate research reporting guidelines and uploaded the relevant EQUATOR Network research reporting checklist(s) and other pertinent material as supplementary files, if applicable.

